# The impact of dance movement interventions on psychological health in older adults without dementia: A systematic review and meta-analysis

**DOI:** 10.1101/2022.11.25.22282727

**Authors:** Odile Podolski, Tim Whitfield, Leah Schaaf, Clara Cornaro, Theresa Köbe, Sabine Koch, Miranka Wirth

## Abstract

Multimodal lifestyle-based interventions that integrate physical, mental and social stimulation could promote mental health and brain resilience against dementia. This meta-analysis examined the efficacy of dance movement interventions (DMI) on psychological health in older adults. Pre-registration was done with PROSPERO (CRD42021265112). PubMed, Web of Science and PsycInfo were searched for randomized controlled trials (RCT) evaluating the effects of DMI (>4 weeks’ duration) on measures of psychological health (primary outcome) and cognitive function (additional outcome) among older adults without dementia (≥55). Data of 13 primary RCT (*n*=943, *n-DMI*=474, *n-control*=469) were synthesized using a random effects meta-analysis with robust variance estimation. DMI had a small positive effect on overall psychological health (*g*=0.31; *95% CI*: [0.09,0.53]; *p*=.01, *I*^*2*^*=*62.55) and a medium effect on general cognitive function (*g*=0.48; *95% CI*: [0.03,0.93], *p*=.04, *I*^*2*^*=*82.45) compared to comparators. None of the primary studies included measures of neuroplasticity. DMI may serve as a multimodal enrichment strategy to promote healthy mental aging. High-quality intervention studies are needed to expand evidence for psychological domains and identify the underlying neurophysiological correlates.

## 1 Introduction

Due to falling fertility rates and longer life expectancy, the global population of older adults continues to grow (United Nations, 2020). As a result, the prevalence of age-related conditions, such as Alzheimer’s disease (AD), has increased. Whilst disease-modifying therapies are emerging, they are not curative, and thus the increased prevalence of AD (the most common cause of dementia) poses tremendous economic as well as social challenges to healthcare systems, patients and their caregivers (Connell et al., 2001; Hurd et al., 2013; Michalowsky et al., 2019). Lifestyle-related physical, cognitive and psychosocial modifiable risk factors contribute to aging processes and the development of age-related conditions (Livingston et al., 2020; Norton et al., 2014). As a consequence, there is an urgent need to establish behavioral strategies that target these risk factors and effectively promote healthy aging, which may ultimately contribute to the prevention of AD (Rosenberg et al., 2020).

Mental or psychological health can be considered a key target for healthy aging and AD prevention strategies. Thereby, it is emphasized that mental health is a psychological state of well-being and psychosocial integration – not just the absence of mental disease (World Health Organization, 2005). Psychological factors including depression and social isolation have been proposed to increase the risk of cognitive impairment and dementia (Livingston et al., 2020), possibly via depleting resilience against brain pathology and accelerating cognitive decline (Marchant and Howard, 2015). Indeed, studies in older adults have indicated that negative affective burden is related to elevated brain AD pathology (Marchant et al., 2020), altered brain functioning (Fredericks et al., 2018; Schwarz et al., 2022), greater cognitive decline and increased risk of AD (Marchant et al., 2020; Terracciano et al., 2017; Wilson et al., 2003). On the other hand, positive psychosocial factors including social engagement, mindfulness, stress resilience and positive thinking styles have been associated with better brain and cognitive health (Demnitz-King et al., 2022; Felix et al., 2020; Whitfield et al., 2021) and reduced risk of dementia (Duffner et al., 2022) in elders.

Lifestyle-based interventions may promote healthy aging by fostering physical, mental, and social well-being, as the three key dimensions of a person’s general health condition (World Health Organization, 2005). To effectively target several risk and/or protective factors at once, it is likely that multimodal or multi-domain interventions will be required, as these effectively combine several beneficial activities (Rosenberg et al., 2020). Studies in animal models have demonstrated far-reaching benefits of environmental enrichment; this typically integrates multimodal stimulation of motor, sensory, cognitive and social processes (Kempermann, 2019). Lifestyle activities suggested to resemble enrichment in humans intrinsically integrate body and mind activities and thus encourage an “*embodied mind in motion*” (Kempermann, 2022). A prime example of an embodied activity are dance movement interventions (DMI), which combine music with movement and mental stimulation embedded in a socially engaging environment. Amongst others DMI may comprise diverse programs and practices including traditional dance, aerobic dance, and dance/movement therapy. The latter is part of the creative arts therapies and incorporates psychotherapeutic factors such as embodiment, creativity and psychosocial processes to promote health and well-being (de Witte, Martina et al., 2021). Overall, DMI thus provide the opportunity to equally influence and balance body and mind, which is proposed to foster healthy aging of physiological, cognitive, and psychological functions (Basso et al., 2021a; Herold et al., 2018) – in agreement with the WHO recommendations for mental health promotion (World Health Organization, 2005).

There is evidence to suggest that engaging in a regular dance activity is associated with a decreased risk of dementia (Verghese et al., 2003). Evidence syntheses have indicated robust positive effects of DMI versus comparators on cognitive (Chan et al., 2020; Hewston et al., 2020; Meng et al., 2020) and physical (Fong Yan et al., 2018; Liu, X. et al., 2021; Mattle et al., 2020) health in the older population. Given the importance of psychological risk factors in the development of cognitive impairment and dementia, advanced knowledge on the impact of DMI on psychological health is needed. According to meta-analyses in a broad sample of studies including younger, middle-aged and older populations, DMI may improve psychological health outcomes including overall, positive and negative psychological functions (Koch et al., 2014; Koch et al., 2019). Yet, evidence on the impact of DMI on the promotion of psychological wellbeing specifically in older adults remains sparse and inconclusive. So far, evidence syntheses in older participants with MCI have suggested that DMI may reduce depression (Wang et al., 2021) and improve quality-of-life (QoL) (Wu et al., 2021), although a different review observed mixed evidence of efficacy across psycho-behavioral outcomes (Liu, C. et al., 2021).

Therefore, the main objective of our study was to conduct a systematic review and meta-analysis of the efficacy of DMI versus comparators for improving psychological health outcomes in older adults without dementia (i.e., clinically normal, subjective cognitive decline [SCD] or mild cognitive impairment [MCI]), using data from randomized controlled trials (RCT). Furthering knowledge on this central health aspect may inspire the design of new-generation DMI, specifically aiming to improve psychological health and wellbeing in the elderly.

## 2 Material and methods

### 2.1 Protocol and registration

This systematic review and meta-analysis was conducted following the Preferred Reporting Items for Systematic Reviews and Meta-Analyses (PRISMA) recommendations (Moher et al., 2009) and was registered with PROSPERO in July 2021 [CRD42021265112].

### 2.2 Criteria for considering studies for this review

#### 2.2.1 Types of studies

RCT were considered eligible for this review. To be included, trials had to clearly describe that participants were allocated via individual or cluster randomization.

#### 2.2.2 Types of participants

The population of interest for this review were adults aged 55 or older without a diagnosis of dementia. Thus, clinically normal older participants or participants with a diagnosis of SCD or MCI were included. To ensure that clinical samples were ascertained according to standardized protocols, diagnostic criteria had to be reported. No restrictions regarding the type of living arrangement were made. Studies with an explicitly defined target population of patients with current or past diagnosis of psychiatric, neurological, inflammatory, and/or other serious medical disorders (e.g., dementia, diabetes, cancer) were excluded.

#### 2.2.3 Types of interventions

We conceptualized DMI as multimodal mind-body activities that involve coordinated movements carried out in synchronization with music and progressing through space. We considered a broad variation of DMI and dance movement styles eligible, as these activities inherently share common basic principles including simultaneous stimulation of multimodal motor, sensory, cognitive, social and emotional processes (Basso et al., 2021a; Koch et al., 2019). Studies had to include a DMI incorporating a pre-specified training program in any style, for example, traditional/ social dance, creative/expressive/ meditative dance, dance/movement therapy or eurythmy. Individual and group interventions were eligible, including both professionally facilitated and self-guided formats. DMI in a virtual or video game format were also eligible for inclusion, although none were eligible following full-text screening. To be included, interventions had to have a minimum duration of 4 weeks. No restrictions for session frequency were applied. Study comparators could be active or passive. Interventions consisting solely of physical exercises in terms of aerobic strength training, coordination training, ergometer training or meditation and breathing exercises were excluded.

#### 2.2.4 Types of outcome measures

##### 2.2.4.1 Primary outcome

The primary outcome of the study was psychological health. This outcome was evaluated through self-reported (subjective) measures related to overall psychological health, including QoL, well-being, depression, anxiety and psychological distress. The outcome measures had to be acquired through validated assessment tools. Due to the limited data on the effects of DMI on psychological health outcomes in the target population, we utilized a broad clustering of psychological outcomes for analyses. This methodological approach was selected in accordance with previous works (Koch et al., 2014; Koch et al., 2019). Firstly, we synthesized all outcome measures across DMI studies to assess overall psychological health.

In addition, we created two subcategories of psychological health, thereafter termed psychological domains. The ‘positive’ domain subsumed measures associated with positive mood and emotions, such as QoL, psychological well-being, and social integration/connectedness. The ‘negative’ domain subsumed measures related to negative mood and emotions, such as depression, anxiety, and psychological distress.

##### 2.2.4.2 Additional outcome

As an additional outcome, we assessed general cognitive function measured through validated neuropsychological assessment tools of general cognitive ability. Given the present focus, only neuropsychological instruments associated with global psychological status or cognitive abilities including the Mini-Mental State Examination (MMSE) (Folstein et al., 1975), the Montreal Cognitive Assessment (MoCA) (Nasreddine et al., 2005), and the Raven’s Progressive Matrices Test (Raven and Raven, 2003) were considered.

### 2.3 Search methods for identification of studies

The following electronic databases were searched: PubMed, Web of Science and PsycInfo. Eligible studies had to have full-text availability and be written in English, and were restricted to peer-reviewed journals and academic publications (dissertations). The initial search was conducted on April 13^th^, 2021 and updated on November 10^th^, 2021.

Key terms and search strings were derived from pre-existing systematic reviews and meta-analyses with related topics (Mattle et al., 2020; Meng et al., 2020; Wu et al., 2019; Zhang et al., 2018) and adapted to the present research question. A combination of keywords, free terms and Medical Subject Headings (MesH) terms was applied across databases. Search strings combined words covering the main elements of the research question including the stems “dance*”, or “music and exercise”, with “older adults”, “psychological health” and interventional terms such as “random*”. Database-specific search strings are provided in the supplementary material.

#### 2.3.1 Selection of studies

After deduplication with the reference manager software EndNote (version X9.3.1) a two-stage screening process was utilized. After a pilot screening, two trained independent reviewers (OP, LS), unblinded to the study authors, screened the identified studies in parallel. At both the title-abstract and full-text stages, studies were evaluated according to the specified inclusion and exclusion criteria. To ensure high standardization and to facilitate screening, a standardized operating procedure (SOP) for the reviewing process was created. Each stage of the screening was completed by comparing decisions of the reviewers. Where no consensus could be reached by discussion, a third independent reviewer (TK, MW) was engaged for the final decision on eligibility. Reasons for exclusion of studies were documented. A PRISMA flow-diagram (Moher et al., 2009) was created for displaying the study selection process. Cohen’s Kappa was calculated to assess inter-rater reliability.

#### 2.3.2 Data extraction and management

Two trained independent reviewers (OP, LS), used a standardized form to extract all study data in parallel. The information extracted covered general information (i.e., study authors, publication date and type, recruitment country, study funding and conflicts of interest), methodological information (i.e., study design and setting, characteristics of participants, descriptions of the intervention and comparator) and outcome information (i.e., outcomes of interest, measurement tools, outcome data reported pre- and post-intervention). Discrepancies between the data extraction forms were discussed until a consensus was reached. Where outcome data were reported using the standard error (SE), a conversion to standard deviation (SD) was performed in order to calculate effect sizes. Studies reporting insufficient data for statistical synthesis were excluded. Where appropriate, study authors were contacted for additional information via e-mail.

#### 2.3.3 Assessment of risk of bias in included studies

For each eligible study, risk of bias was assessed during data extraction with the Risk of Bias 2 tool (Sterne et al., 2019) of the Cochrane Collaboration. Each study was rated according to an intention-to-treat approach and in respect to the primary outcome (i.e., psychological health) only. Each of the following domains (D) was rated in parallel by two independent reviewers (OP, LS) as either “high risk”, “some concerns” or “low risk”: (D1) bias arising from the randomization process; (D2) bias due to deviations from intended interventions; (D3) bias due to missing outcome data; (D4) bias in measurement of the outcome; (D5) bias in selection of the reported result. The overall bias judgement is reached by an algorithm based on the answers given to several signaling questions within each domain. Disagreement was resolved by discussion until consensus was reached.

#### 2.3.4 Assessment of heterogeneity

The parameter *I*^*2*^ was used to assess heterogeneity within each model. The *I*^*2*^ statistic represents the percentage of variability in effect sizes not attributable to sampling error. Interpretation followed a previously described rule of thumb (Higgins et al., 2003) with *I*^*2*^ = 25% meaning low heterogeneity, *I*^*2*^ = 50% moderate heterogeneity and *I*^*2*^ = 75% substantial heterogeneity.

#### 2.3.5 Assessment of reporting bias

A funnel plot created in R using the “metafor” package (version 3.0-2) was used to graph the observed effect sizes against standard errors. Interpretation and assessment of asymmetry was supported by employing Egger’s regression test (Egger et al., 1997), indicating publication bias through a significant result.

### 2.4 Data synthesis

For the calculation of effect sizes for controlled designs with pre- and post-test scores, the standardized mean difference (SMD; with 95% confidence intervals (*CI*)) with a bias correction (*g*) was used (Borenstein et al., 2021; Hedges, 1981; Morris, 2008). Those outcome assessments with lower scores indicating an improvement after DMI were inverted, such that higher values reflected improvement or better performance for any given outcome. The calculations (see supplementary material for detailed description and formula) were applied and implemented using the free statistical computation software R (version 4.0.3) for each outcome score to determine effect sizes.

#### 2.4.1 Accounting for dependencies

As defined in the inclusion criteria, the outcomes of interest could be assessed via multiple assessment tools within one study. Incorporating more than one outcome measure per study violates the assumption of independent effect sizes of common meta-analysis models (Tanner-Smith et al., 2016). To overcome this problem, typically only one effect size or an average effect size per study is selected. These procedures result in loss of information and should therefore be avoided (Matt and Cook, 1994).

To adequately account for dependencies in outcome measures within studies, we used a random effects meta-analysis with robust variance estimation (RVE) (Hedges et al., 2010). Modelling was implemented through the “robumeta” package (version 2.0) in R with a correction for small-samples (Fisher and Tipton, 2015). The package provides a default setting of *rho* at 0.8 for the assumed correlation of outcomes within studies. Sensitivity analyses varying *rho* from 0–1 were run to investigate the possible impact on *Tau*^*2*^ as estimator of variance.

Results were defined as being significant with a *p*-value of < .05. Where degrees of freedom fall below 4, *p*-values for RVE meta-analytic estimates are considered unreliable (Fisher and Tipton, 2015), and are therefore not reported. The following rubric was used to interpret effect sizes: *g* ≥ 0.2 (small), *g* ≥ 0.5 (medium), and *g* ≥ 0.8 (large) (Cohen, 2013). Forest plots were created in R to visualize results.

#### 2.4.2 Subgroup and outlier analysis

As the number of included studies was expected to be small, no meta-regression analyses were planned. Exploratory subgroup analyses were conducted to further investigate the effects of DMI. Subgroup analyses should not be collectively regarded as direct comparisons, but rather to inform future research. Subgroup analyses stratified results by: (i) Cognitive status (clinically normal; MCI), (ii) Control group type (active; passive), (iii) Intervention duration (≤ 16 weeks; > 16 weeks), (iv) Intervention type (creative arts therapy; traditional/ballroom/aerobic dance). Due to the limited data and heterogeneity the subgroup analyses need to be interpreted with caution.

Lastly, an exploratory post-hoc outlier analysis was carried out. For this, effect sizes of outcomes with *CIs* outside the boundaries of the pooled effect *CI* across studies were classified as outliers (Viechtbauer and Cheung, 2010). Additionally, when in doubt of methodological quality or where high probability of bias occurred, individual studies were excluded to explore influence on effects. Subsequently, analyses described before were repeated excluding studies classified as outliers.

## 3 Results

### 3.1 Study selection

The initial systematic search across the three databases in April 2021 yielded a total of 640 records. After deduplication, 484 records remained for title-abstract screening. Two independent reviewers (OP, LS) screened the title-abstracts with an inter-rater reliability (unweighted Cohen’s kappa) of 0.80 (95% *CI* [0.73, 0.88]). Studies meeting eligibility criteria were full-text screened by OP and LS in parallel reaching high agreement with an unweighted kappa of 0.93 (95% *CI* [0.84, 1.02]). During the process of data extraction, 15 studies were excluded. Reasons for exclusion were documented. Study selection resulted in a final number of 12 primary studies (see Figure 1 for PRISMA flowchart). An additional primary study was included after the search update in November 2021, resulting in a total of 13 studies.

**Figure 1.**
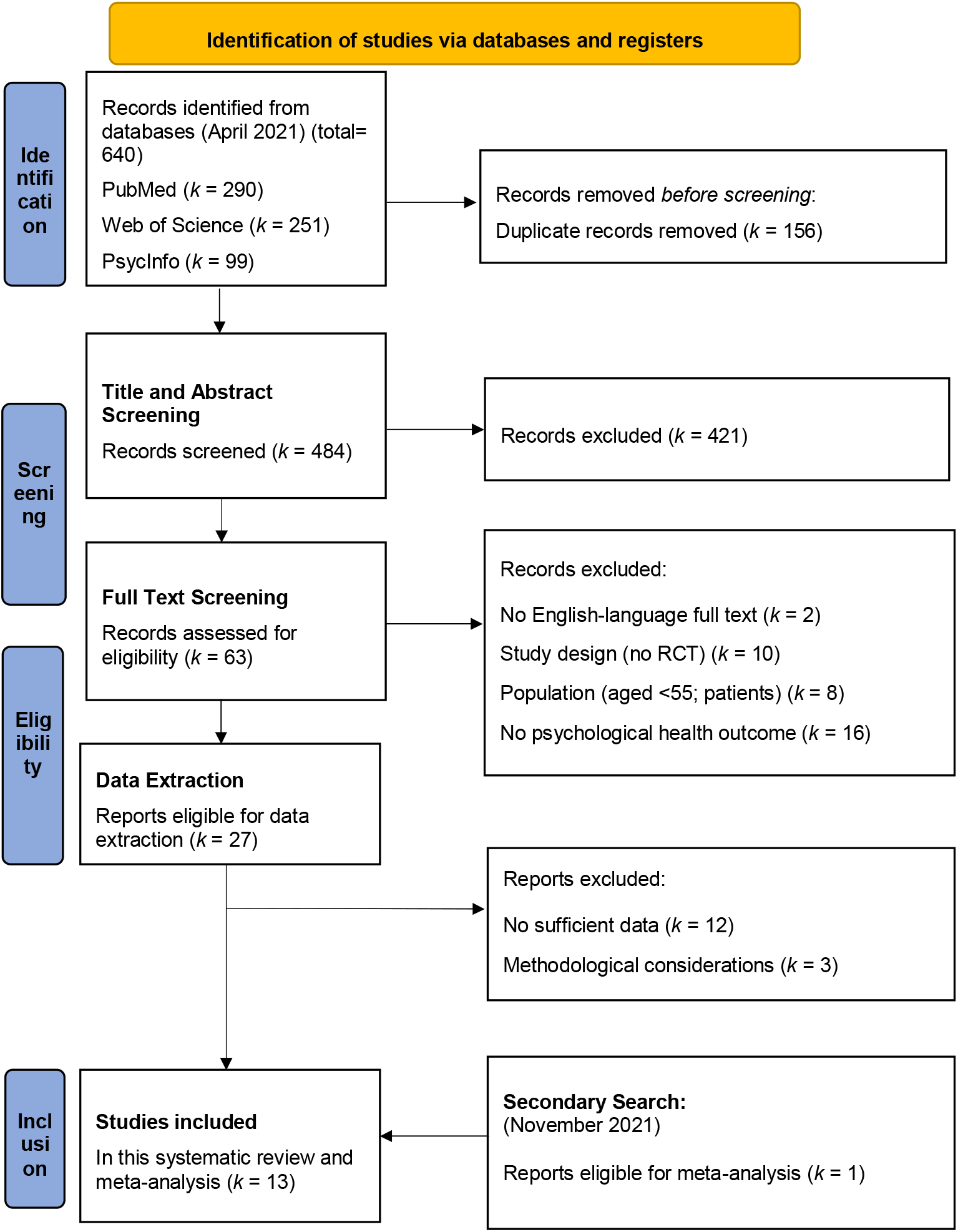
PRISMA flowchart. **Key**. *k*, Number of studies; *RCT*, randomized-controlled trial.

### 3.2 Study characteristics

An overview of study characteristics is provided in Table 1. Publication years ranged from 2009 to 2020. Seven studies (54%) were conducted in Europe, four studies (31%) in Asia and one study (7%) in Canada and Brazil, respectively. Twelve peer-reviewed journal articles and one dissertation (Alves, 2014) were included. One study (Hars et al., 2014) was a secondary analysis of an RCT reported elsewhere (Trombetti et al., 2011). Psychological health measures were a primary outcome in only one study (Liao et al., 2018) and were otherwise included as secondary outcomes. The majority of studies used cognitive function (*k* = 8; 62%) or physical function (*k* = 4; 31%) as the primary outcome.

**Table 1.**
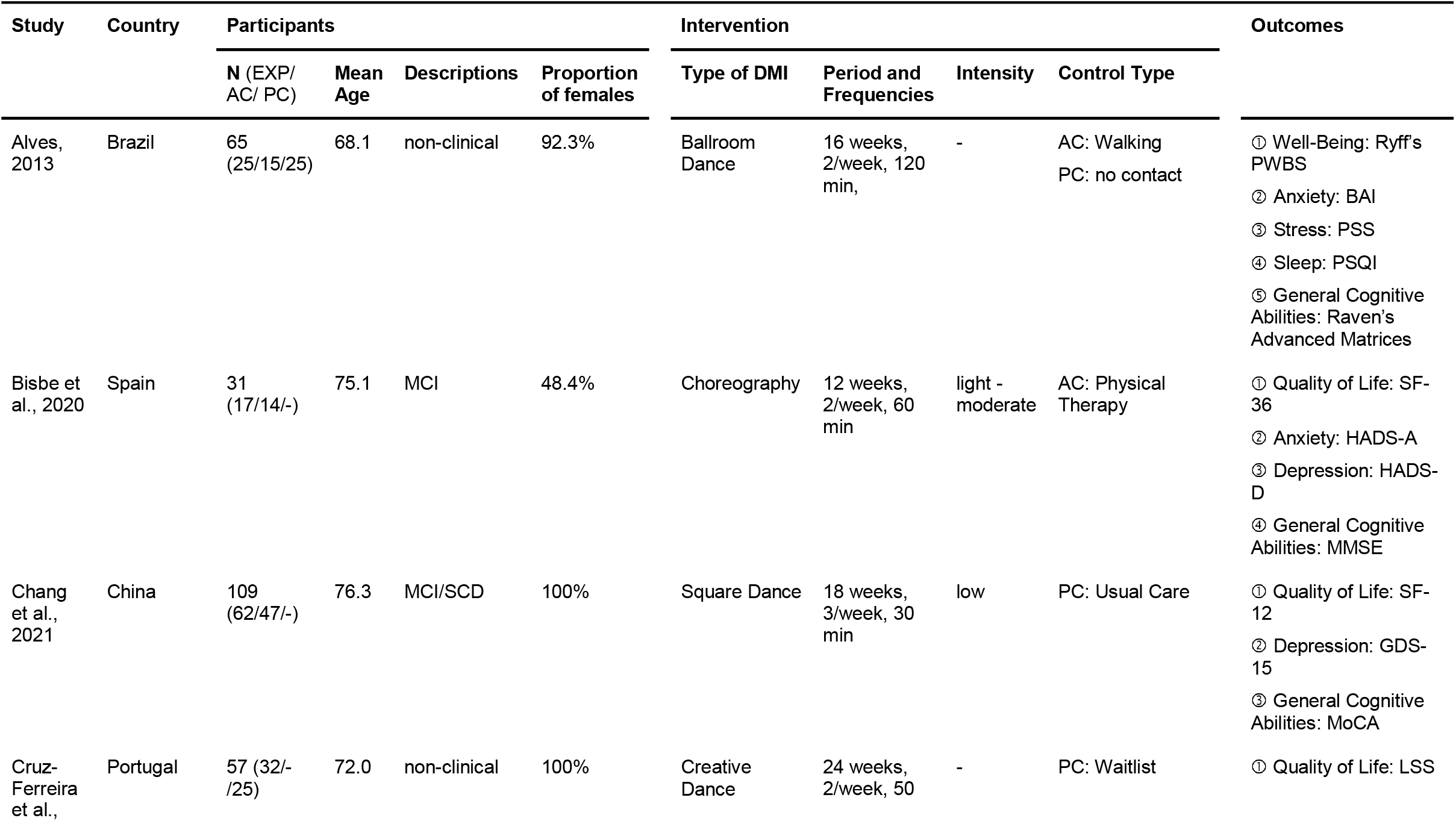
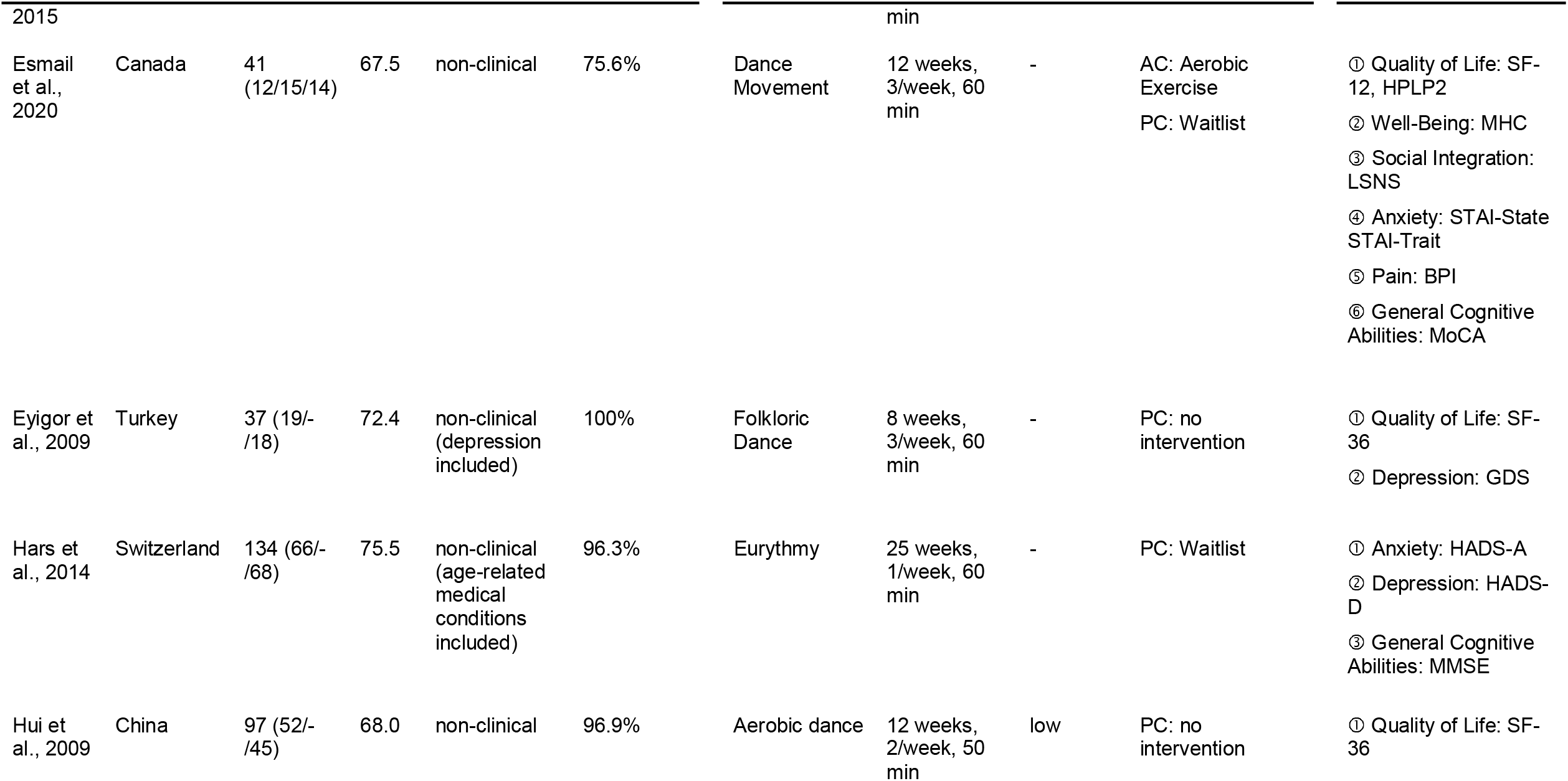
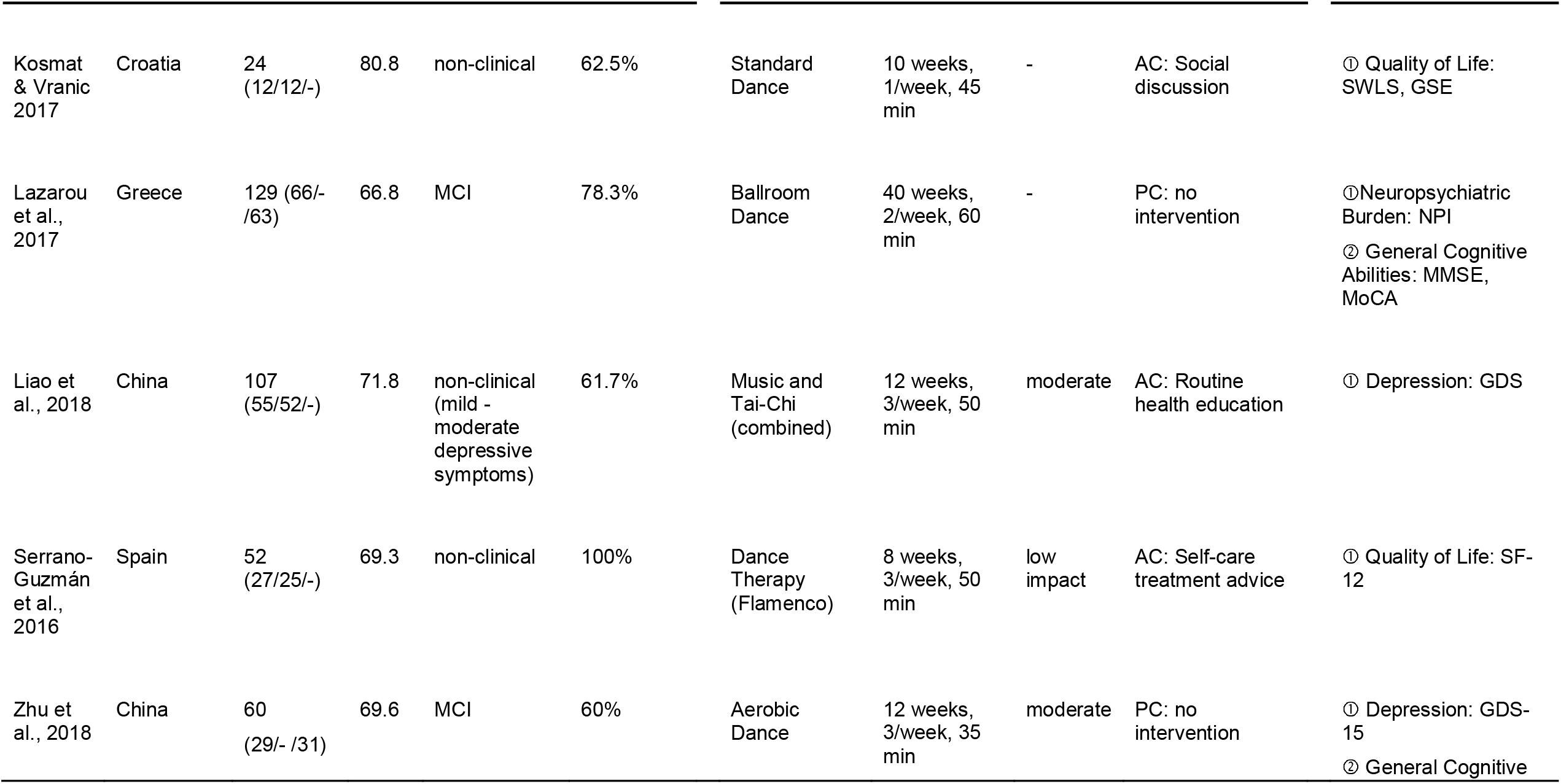
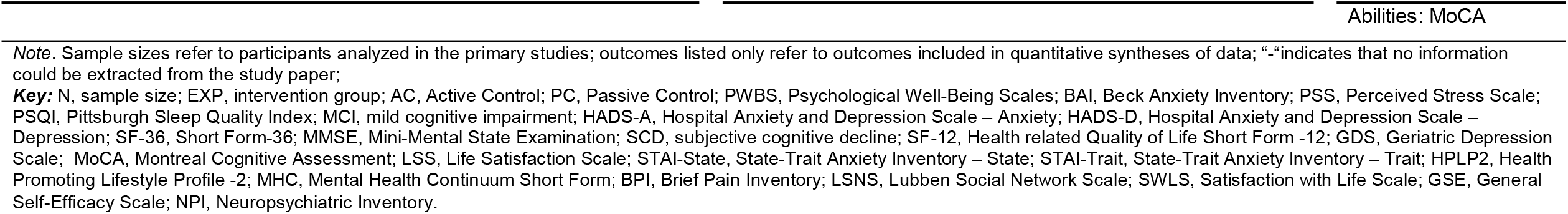
Characteristics of included studies.

Most of the studies employed randomization at the individual level (*k* = 11; 85%); one study used cluster randomization (Liao et al., 2018). Another study (Hui et al., 2009) divided participants into two groups prior to randomization to maintain social peers. Two trials applied a three-armed design with two control group types (Alves, 2014; Esmail et al., 2020). Four studies (31%) reported several time points of assessment during the intervention (Chang et al., 2021; Esmail et al., 2020; Liao et al., 2018; Zhu et al., 2018). Only one study (Kosmat and Vranic, 2017) reported a follow-up assessment beyond the end of intervention.

### 3.3 Participant characteristics

This review included data of 943 participants (*n-DMI* = 474, *n-control* = 469). Sample sizes varied from 24 to 134 participants per study. Participants enrolled in the studies had an average age of 72 years. The average percentage of female participants in the reported samples was 82.5%. Four studies (31%) included female participants only. Eight studies (62%) recruited healthy older adults; one study included age-related health conditions (Hars et al., 2014). The remaining four studies (31%) investigated participants with a confirmed diagnosis of MCI (Bisbe et al., 2020; Chang et al., 2021; Lazarou et al., 2017; Zhu et al., 2018). None of the primary studies explicitly investigated older adults with SCD. Five studies (38%) provided information on education in years with an average of 11 years of education.

Overall, it was reported that included participants neither had experience of dance nor engaged in similar activities before starting the trial. Of note, 10 studies (77%) reported specific assessments of activity levels or whether participants engaged regularly in physical exercise prior to intervention. Five studies (38%) examined a sample defined as inactive (Esmail et al., 2020) or (moderately) sedentary (Alves, 2014; Bisbe et al., 2020; Chang et al., 2021; Serrano-Guzmán et al., 2016), whereas two studies examined participants with an active lifestyle prior to intervention (Eyigor et al., 2009; Liao et al., 2018).

### 3.4 Intervention characteristics

#### 3.4.1 Intervention type / types of DMI

A comprehensive overview of intervention characteristics is provided in Supplementary Table S1. Types of DMI varied across the studies. Five studies (38%) included standard or traditional dance (e.g., ballroom dance, folkloric dance) (Alves, 2014; Chang et al., 2021; Eyigor et al., 2009; Kosmat and Vranic, 2017; Lazarou et al., 2017). Three studies (23%) reported aerobic dance (Bisbe et al., 2020; Hui et al., 2009; Zhu et al., 2018). Five studies (38%) were categorizable as evaluating creative arts therapies (de Witte, Martina et al., 2021). Those included interventions designed according to standards of the American Dance Therapy Association (Esmail et al., 2020; Serrano-Guzman et al., 2016) and a creative dance program (Cruz-Ferreira et al., 2015). Hars and colleagues (2014) employed eurythmy to apply multitask practices to piano music and Liao and colleagues (2018) used Tai-Chi movements combined with music.

#### 3.4.2 Intervention period, duration and frequency

The median intervention duration was 12 weeks (range: 8 to 40 weeks). Nine studies (69%) had a length of 16 weeks or below (Alves, 2014; Bisbe et al., 2020; Esmail et al., 2020; Eyigor et al., 2009; Hui et al., 2009; Kosmat and Vranic, 2017; Liao et al., 2018; Serrano-Guzmán et al., 2016; Zhu et al., 2018) four studies (31%) were conducted over 18 weeks or more (Chang et al., 2021; Cruz-Ferreira et al., 2015; Hars et al., 2014; Lazarou et al., 2017). Frequencies of DMI sessions varied from once per week to three times per week, while individual session durations lasted from 35 to 120 minutes. The training dose ranged from a minimum of ten DMI sessions with 45 minutes’ duration (Kosmat and Vranic, 2017) to a maximum dose of 80 sessions of 60 minutes (Lazarou et al., 2017).

Across studies, the structure of all DMI sessions comprised a warm-up, followed by the training phase and subsequently a cool-down or stretching phase. Notably, one study (Eyigor et al., 2009) required participants to walk for approximately 30 minutes twice a week in addition to the DMI. Information regarding the intensity of DMI sessions was presented in six studies (46%). Three studies (23%) had low intensity (Chang et al., 2021; Hui et al., 2009; Serrano-Guzmán et al., 2016) and three had (23%) moderate intensity sessions (Bisbe et al., 2020; Liao et al., 2018; Zhu et al., 2018). One study ensured that the target intensity was achieved by measuring the individual heart rates during training (Zhu et al., 2018).

#### 3.4.3 Intervention setting

All studies performed DMI in a group setting. In three studies (15%) the DMI was designed to encourage social interaction between pairs or group members (Alves, 2014; Cruz-Ferreira et al., 2015; Eyigor et al., 2009). For the remaining interventions each person performed the exercises individually. Eight studies (62%) were conducted at local healthcare centers, hospitals or similar public facilities. One DMI took place in an outdoor setting (Chang et al., 2021). Most of the interventions (*k* = 11; 85%) were led by a certified or experienced (dance) instructor or a facilitator specially trained for the DMI program. Two studies (15%) (Liao et al., 2018; Serrano-Guzman et al., 2016) did not provide information on instructor/facilitator qualification.

#### 3.4.4 Intervention adherence

Nine studies (69%) provided data on adherence for the DMI, mainly reported as the proportion of classes or courses attended. Zhu and colleagues (2018) reported the median number of sessions attended. Across studies, DMI attendance rates ranged from 79% to 100%; five studies (38%) reported adherence of 90% or more (Alves, 2014; Bisbe et al., 2020; Esmail et al., 2020; Hui et al., 2009; Serrano-Guzman et al., 2016). Across studies, the proportion of individuals who discontinued study participation ranged from 0 to 48% as assessed for intervention and control groups. Two studies reported that all participants completed the trial (Cruz-Ferreira et al., 2015; Serrano-Guzman et al., 2016). Three studies (23%) were unclear regarding the proportion of participants who dropped out.

#### 3.4.5 Control condition

Eleven studies (85%) included a single comparator condition (active or passive), whereas two studies included both an active and passive control group (Alves, 2014; Esmail et al., 2020). Of the eleven studies employing one comparator, seven evaluated DMI against a passive control group of waitlist (*k* = 2) (Cruz-Ferreira et al., 2015; Hars et al., 2014), usual care (*k* = 1) (Chang et al., 2021), or no intervention conditions (*k* = 4) (Eyigor et al., 2009; Hui et al., 2009; Lazarou et al., 2017; Zhu et al., 2018). Four studies utilized an active comparator, including physical therapy (Bisbe et al., 2020), discussion groups (Kosmat and Vranic, 2017), routine health education (Liao et al., 2018) and self-care advice (Serrano-Guzmán et al., 2016).

### 3.5 Neurophysiological measures

None of the primary studies has evaluated measures of neuroplasticity potentially underlying the effect of DMI on psychological health outcomes.

### 3.6 Risk of bias

Methodological quality of the primary studies varied. A visual summary of risk of bias ratings is provided in Figure 2. Overall risk of bias was rated as being at “high risk” in nine studies (69%), while four studies (31%) were rated as having “some concerns”; no study was rated as “low risk”. Please see the supplementary material for domain-specific risk of bias ratings.

**Figure 2.**
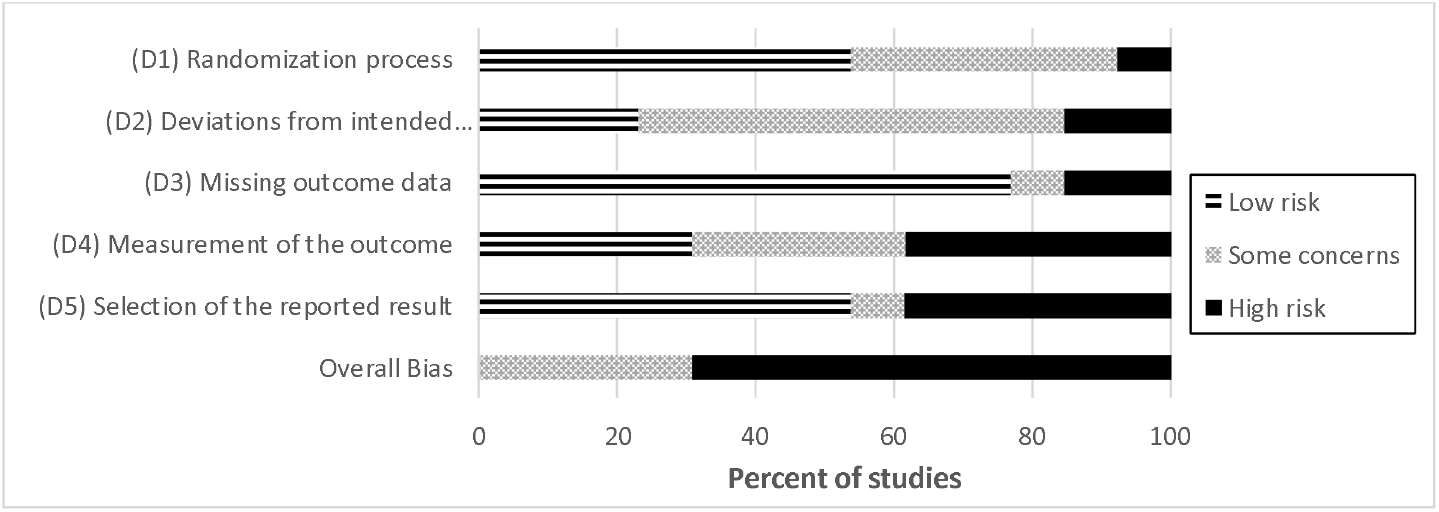
Summary of Risk of Bias Rating according to domains. Note. The risk of bias graph presents ratings for the 13 included studies rated regarding an intention to treat approach.

### 3.7 Publication bias

The distribution of effect sizes and SEs was visualized using a funnel plot (see supplementary material). Whilst a degree of asymmetry was apparent on the funnel plot, an Egger’s test found no evidence of publication bias (intercept, -0.63; *p* = .53; *CI* 95% [-0.12, 1.20]).

### 3.8 Quantitative synthesis of results

#### 3.8.1 Primary Outcome: Psychological health

Across studies, DMI outperformed comparators for overall psychological health (*g* = 0.31, *p* = .01; see Table 2). The result was based on the pooled data of psychological health outcomes (*k* = 13, see forest plot Figure 3) and compared to passive control (*k* = 9), where applicable (for studies utilizing both passive and active comparators). The *I*^*2*^ statistic indicated a moderate amount of heterogeneity among the included studies. Sensitivity analyses varying the parameter *rho* did not change the value of *Tau*^*2*^, representing robust variance estimation against dependencies.

**Table 2.** Meta-analyses comparing DMI to passive control type (where applicable) for psychological health outcomes.

|  | Construct | k (NES) | ES | 95% CI | SE | df | p-value | I<sup>2</sup>% |
| --- | --- | --- | --- | --- | --- | --- | --- | --- |
| Overall psycho-logical health | all combined | 13 (42) | 0.31 | [0.09, 0.53] | 0.10 | 11 | .01 | 62.55 |
| Positive domain | all combined | 10 (29) | 0.26 | [-0.07, 0.59] | 0.15 | 9 | .12 | 68.18 |
|  | Quality of life | 10 (22) | 0.31 | [-0.08, 0.71] | 0.20 | 9 | .12 | 74.03 |
|  | Social Integration | 3 (3) | 0.85 | [-1.59, 3.29] | 0.57 | 2* | * | 85.17 |
| Negative domain | all combined | 8 (13) | 0.30 | [-0.05, 0.65] | 0.15 | 7 | .08 | 66.77 |
|  | Anxiety | 4 (5) | 0.16 | [-0.71, 1.03] | 0.27 | 3* | * | 70.67 |
|  | Depression | 5 (5) | 0.22 | [-0.25, 0.68] | 0.16 | 4 | .26 | 60.81 |
| General cognitive abilities | all combined | 7 (8) | 0.48 | [0.03, 0.93] | 0.18 | 6 | .04 | 82.45 |

**Key:** DMI = Dance Movement Interventions; k, number of studies; NES, Number of Effect Sizes; ES, Effect size; CI, Confidence interval; SE, Standard Error; df, Degrees of freedom.
\*Where df < 4, p-values are unreliable and are thus not reported.

**Figure 3.**
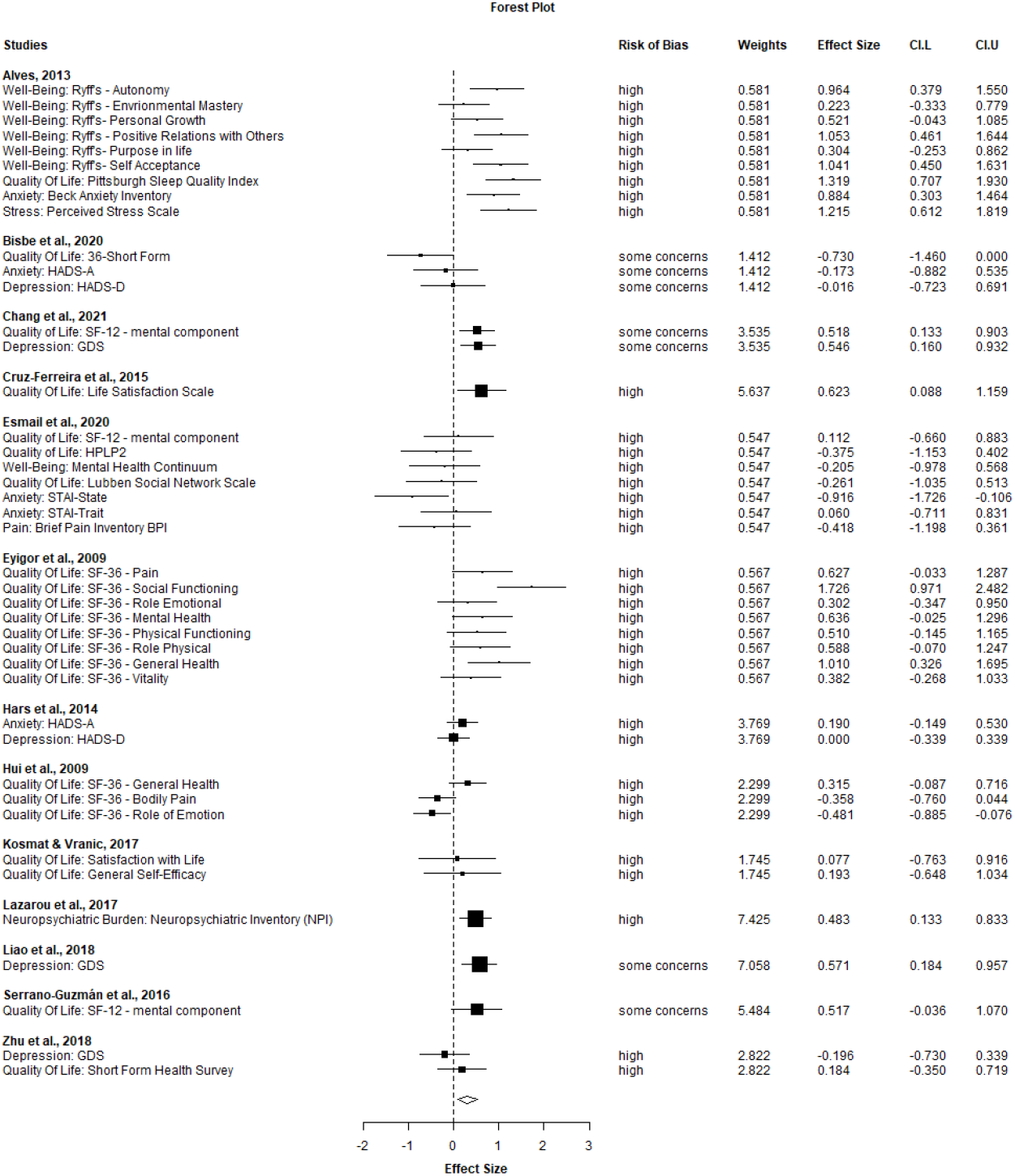
Forest Plot of effect sizes of DMI compared to passive control type (where applicable otherwise active control groups were used as comparator) on psychological health outcomes for individual studies and outcome measures. *Note*. Effect sizes are presented for each study according to the construct measured and assessment tools used, respectively. Values on the left side of the null line favor control condition and values on the right side favor DMI. **Key:** CI.L, Confidence interval lower bound; CI.U, Confidence interval upper bound, HADS-A/D, Hospital Anxiety and Depression Scale; HPLP-2, Health Promoting Lifestyle Profile 2; STAI, State Trait Anxiety Inventory; GDS, Geriatric Depression Scale.

Separation of psychological health outcomes into positive and negative domains revealed non-significant trends favoring DMI over comparators (see Table 2) with small improvements in both the positive (combined: *g* = 0.26, *p* = .12) and negative domains (combined: *g* = 0.30, *p* = .08). Notably, there was a large positive effect of DMI on social integration (*g* = 0.85), however, degrees of freedom fell below four resulting in a low reliability of effect size. Both positive and negative domains showed moderate to substantial (61 - 85%) heterogeneity. Sensitivity analysis varying *rho* did not impact variance estimation. Forest plots are provided in the supplementary material.

#### 3.8.2 Additional Outcome: Cognitive function

As expected, DMI outperformed comparators for general cognitive abilities, with a medium effect size (*g* = 0.48, *p* = .04; see Table 2). The observed heterogeneity was substantial (*I*^*2*^ > 80%). Excluding the Raven’s Matrices Scale in a post-hoc analysis revealed a similar, albeit non-significant effect size, while heterogeneity remained substantial (*g* = 0.44; 95% *CI* [-0.11, 0,98]; *p* = 0.09; *I*^*2*^ = 85.13)

#### 3.8.3 Exploratory subgroup analysis

Results of the exploratory subgroup analysis are given in Table 3. Significant subgroup results on overall psychological health were found for studies that incorporated clinically normal participants and for studies using passive control conditions. Above that, studies with a longer intervention duration (> 16 weeks) showed a medium effect size, however, degrees of freedom fell below four resulting in a low reliability of effect size. For the type of DMI, effect sizes were similar for the subgroups of creative arts therapy and aerobic dance (including traditional, ballroom and aerobics), both showing positive trend level effects on psychological health. The amount of heterogeneity ranged from small to moderate across subgroups (*I*^*2*^_*min*_ = 35%; *I*^*2*^_*max*_ = 70%).

**Table 3.** Effect sizes on overall psychological health according to exploratory subgroup analyses comparing DMI to passive control type (where applicable).

| Subgroup |  | k (N ES) | ES | 95% CI | SE | df | p-value | I<sup>2</sup> % |
| --- | --- | --- | --- | --- | --- | --- | --- | --- |
| Cognitive status | MCI | 4 (8) | 0.26 | [-0.33, 0.84] | 0.18 | 3* | * | 59.33 |
|  | Clinically normal | 9 (34) | 0.34 | [0.03, 0.65] | 0.13 | 8 | .03 | 67.22 |
| Intervention duration | ≤ 16 weeks | 9 (36) | 0.24 | [-0.10, 0.58] | 0.15 | 8 | .15 | 69.50 |
|  | > 16 weeks | 4 (6) | 0.40 | [-0.01, 0.81] | 0.16 | 3* | * | 35.46 |
| Control group type | Active control | 6 (23) | 0.31 | [-0.13, 0.76] | 0.17 | 5 | .13 | 54.79 |
|  | Passive control | 9 (35) | 0.32 | [0.02, 0.61] | 0.13 | 8 | .04 | 69.14 |
| Intervention type | CAT | 5 (12) | 0.34 | [-0.08, 0.76] | 0.15 | 4 | .08 | 50.97 |
|  | Aerobic dance | 8 (30) | 0.29 | [-0.06, 0.64] | 0.15 | 7 | .09 | 70.35 |

**Key.** DMI, Dance Movement Intervention; k, number of studies; N ES, Number of Effect Sizes; ES, Effect size; CI, Confidence interval; SE, Standard Error; df, Degrees of freedom; MCI, Mild cognitive impairment; CAT, Creative Arts Therapy.
\*Where df <4, p-values are unreliable and are thus not reported.

#### 3.8.4 Outlier analysis

Results of the outlier-adjusted analyses are given in the supplementary material. Exclusion of six individual effect sizes corroborated our results for overall psychological health (*g* = 0.37, *p* < .01) as well as positive (*g* = 0.42, *p* < .01) and negative (*g* = 0.31, *p* = 0.05) domains. Heterogeneity assessed with the *I*^*2*^ -statistic nominally decreased to a small amount, justifying the exclusion of the respective scales. Post-hoc exclusion of one study (Hui et al., 2009) due to methodological considerations further substantiated our results.

## 4 Discussion

### 4.1 Summary of main findings

This systematic review and meta-analysis investigated the effects of DMI on psychological health outcomes in older adults without dementia. In total, 13 primary RCT with 943 participants (*n-DMI* = 474, *n-control* = 469) were synthesized. Analyses revealed a small positive effect of DMI on psychological health compared to control conditions, while likewise improving cognitive function, assessed as an additional outcome. Our findings predominantly relate to clinically normal older adults, while four studies included participants with MCI. The DMI varied in type, frequency and duration resulting in high variability across RCT. Exploration of subgroups suggested that longer duration might foster effectiveness of the intervention on psychological health. Overall, our study indicates that DMI can be considered a promising tool to promote healthy mental aging and may thereby contribute to the prevention of AD among older adults. High-quality intervention studies are needed to expand evidence for psychological domains and determine the underlying neurophysiological correlates.

### 4.2 DMI and psychological health as primary outcome

We demonstrate that DMI have a small positive effect on overall psychological health (*g* = 0.31) compared to control conditions. Re-running analyses after exclusion of outliers corroborated the effect (*g* = 0.37). The finding converges with the previous meta-analysis across different age groups by Koch and colleagues (2019), showing an overall positive effect of DMI on combined psychological health outcomes with a medium effect size. Further evaluating the positive psychological domain, we report a non-significant trend favoring DMI over comparators with a small effect size (*g* = 0.26); this effect was substantiated after exclusion of outliers (*g* = 0.42). The result is in line with evidence synthesis by Wu and colleagues (2021), reporting a significant positive effect of DMI on QoL in persons with MCI, which was however not found by others (Liu, C. et al., 2021). Previous syntheses of DMI studies have demonstrated improvements in QoL, subjective well-being, positive affect and interpersonal skills following DMI compared to comparators (Koch et al., 2014; Koch et al., 2019).

Less clear findings are reported for the negative psychological domain (Alpert et al., 2009; Jeon et al., 2005). We found a non-significant trend favoring DMI over comparators in the negative psychological domain with a small effect size (*g* = 0.30). The most promising evidence for DMI in this domain mainly relates to clinical populations including patients with dementia, showing a reduction of depressive symptoms in response to DMI (Crumbie et al., 2015; Ho et al., 2020). Evidence in a broader sample showed medium effects with high heterogeneity in the reduction of depression and anxiety (Koch et al., 2019). Interestingly, we observed a large positive effect of DMI on social integration (*g* = 0.85). Despite limited reliability due to the small number of studies, this finding deserves further investigation since social isolation is suggested as a risk factor of AD (Livingston et al., 2020). Recent pilot trials have reported enhanced social connectedness after a single online dance session in adults aged ≥ 18 years (Rugh et al., 2022) and a reduction in feelings of social isolation in older adults with SCD/MCI after a 3-month movement program based on principles of creative arts therapy (Chao et al., 2021). DMI might thus be expected to foster social participation in older adults, shown to be protective against dementia (Duffner et al., 2022).

### 4.3 DMI and cognitive function as additional outcome

Our meta-analysis further shows a positive impact of DMI on cognitive function compared to comparators with a medium effect size (*g* = 0.48). In the present study, we analyzed general cognitive abilities as additional outcome. Therefore, the effect is limited to the data reported in the primary trials, as we did not conduct an independent search for this main outcome. Hence, there may be a bias in the data selection. The present result replicates prior meta-analytical studies, showing robust beneficial effects of DMI on several cognitive domains including executive functions (Noguera et al., 2020), memory (Meng et al., 2020) and global cognition (Hewston et al., 2020) in clinically normal older adults and in participants with MCI (Chan et al., 2020; Zhu et al., 2020).

### 4.4 Putative neurophysiological mechanisms underlying psychological benefits

None of the synthesized studies has evaluated neurophysiological markers, thus limiting our understanding of candidate mechanisms underlying the psychological benefits of DMI. Other studies have however suggested that DMI could activate beneficial plasticity at multiple neurophysiological levels, some of which are involved in emotion regulation and processing. According to comprehensive reviews, dance could promote plasticity in functional and structural brain networks (Basso et al., 2021b; Teixeira-Machado et al., 2019). Recent interventional studies in older adults confirmed partially increased functional connectivity in higher-order systems, including the default mode and fronto-parietal networks, in response to 3-, 4- and 6-months dance-based programs, respectively (Balazova et al., 2021; Balbim et al., 2021; Chao et al., 2021). Other studies in similar populations have demonstrated improvements after DMI compared to comparators in white matter microstructural integrity of the fornix (Burzynska et al., 2017) and gray matter morphology. The latter encompassed distributed brain regions including the cingulate cortex, insula and the corpus callosum (after 6 months) along with the hippocampus (after 18 months) (Rehfeld et al., 2017), although another study reported no change in hippocampal volume after a 4-month dance program (Guzman et al., 2021). Interestingly, one trial showed reduced salivary cortisol concentrations during the cortisol awakening response, as a marker of chronic stress, after a 3-month dance/movement program compared to aerobic exercise (Vrinceanu et al., 2019). Whether and how these physiological changes are associated with psychological changes in response to DMI will need further investigation in prospective RCT including sensitive measures of neuroplasticity in the circuitry of emotion regulation.

### 4.5 Recommendations for future studies

Our results support the view that DMI could act as a promising tool to promote mental health in the older population. It is thus important to determine the best possible configuration of DMI (i.e., duration, intensity, type and setting) to enhance and sustain these positive effects. The reported adherence rates in the primary studies were high, suggesting that the older participants were motivated and compliant. This fact may facilitate transfer of this lifestyle activity into everyday life, which may help to sustain benefits of the intervention (Herold et al., 2018). Our exploratory subgroup analysis implied that a longer intervention duration (> 16 weeks) might be more effective to improve psychosocial outcomes, as suggested previously (Wu et al., 2021). Above this, the type of DMI could be an important factor. When we separated the interventions into creative arts therapies or aerobic dance, similar effect sizes were yielded in our small subgroups. Notably, Koch and colleagues (2019) showed that dance/movement therapy positively influenced psychological health in both positive and negative domains. While interventions based on creative arts therapies could be particularly suitable to promote and sustain psychological benefits (de Witte, M. et al., 2021), only a few primary studies have incorporated these principles. Overall, more research is needed to draw valid conclusions on the effectiveness of different DMI configurations on psychological health outcomes.

The present findings clearly indicate that high-quality trials are required to further evaluate the effects of DMI on psychosocial measures as designated primary outcomes. Across the included studies, we observed high statistical heterogeneity and risk of bias across studies, as documented previously (Koch et al., 2019). Whilst a relatively high proportion of studies were rated at high overall risk of bias, this can be attributed to the assessment algorithm of the RoB 2 tool, which downgrades studies with self-reported outcomes. Another explanation relates to the nature of non-pharmacological interventions, where blinding is only possible to a certain extent. To distinguish between nonspecific and specific intervention effects, active control groups (receiving an alternate treatment) are needed. Furthermore, we found that data regarding long-term effects of DMI were scarce, indicating the necessity of follow-up measurements in future trials. We also noticed that the vast majority of participants were female. This should be considered in trial protocols, by tailoring recruitment strategies to attract more male participants. Lastly, neurophysiological correlates are rarely assessed in existing trials and need to be included in prospective studies. This is in line with a bidirectional view of the body (including the brain) and mind as integrated and interacting parts of an adaptive system that should be the fundament of future trials. Well-designed RCT will foster understanding of the proposed mind-and-body benefits of DMI.

### 4.6 Synopsis and outlook

It has been recognized that dance activities could serve as an embodied prevention strategy to promote brain reserve and resilience against dementia (Kempermann, 2022). As a multimodal intervention, dance activities encourage a complex enrichment through the simultaneous activation and synchronization of motor, sensory, cognitive, social and emotional processes (Basso et al., 2021a). The inherent integration and synergy of body and mind is proposed to act as a protective factor against age- and disease-related decline. In this light, the psychological benefits of DMI, as highlighted by our study, may be particularly important for populations at increased risk of AD. Those include older adults diagnosed with SCD, a population characterized by elevated negative psychological burden and/or altered functioning across brain networks subserving emotion regulation and self-referential thought (Perrotin et al., 2017; Sannemann et al., 2020; Schwarz et al., 2022). Yet, none of the synthesized studies has specifically investigating participants with SCD, as a designated target group for early non-pharmacological intervention strategies to enhance brain resources or reduce incipient symptoms (Jessen et al., 2014; Smart et al., 2017).

Based on the current and previous findings, we perceive an overall need to develop and evaluate new-generation DMI. As guiding principles, such intervention programs should 1. specifically aim to promote mental health and wellbeing in the elderly, 2. be easily accessible by implementation into onsite/online settings (Rugh et al., 2022) and 3. tailored to the specific needs of clinical/nonclinical older populations (Chao et al., 2021; Esmail et al., 2020). For example, the incorporation of dance/movement therapy, mindful movement practices and psychotherapeutic factors into targeted DMI programs for older people might promote mental capacities, such as creativity, positive thinking and mindfulness, that have far-reaching benefits for the brain, cognitive and mental health in older age (e.g., Chételat et al., 2022; Demnitz-King et al., 2022; Marchant et al., 2021; Whitfield et al., 2021). Mastering new skills including creative dance movements could improve self-efficacy in elders, which in turn may increase sense of mastery and autonomy as essential coping factors with regard to age-related conditions (Fancourt and Finn, 2019; Gerino et al., 2017). Together such strategies could pave the road towards effective and accessible multimodal interventions with high adherence to promote healthy mental ageing and prevent AD in the long term.

### 4.7 Strengths and limitations

To our knowledge, this is the first systematic review and meta-analysis investigating the effect of DMI on psychological health outcomes in older adults without dementia. The review was strictly conducted according to a pre-specified open source protocol, as stipulated by relevant guidelines (Moher et al., 2009). Screening, risk of bias assessment and data extraction were performed in parallel by two independent reviewers to ensure the accuracy of the data. Bias rating was conducted with the revised and updated Risk of Bias 2 tool, which was designed to assess bias more accurately in light of developments in the field (Sterne et al., 2019). Statistical analyses and data synthesis followed approved methodology applied in the field (Borenstein et al., 2021). Additionally, multiple effect sizes of studies were considered in our meta-analytic approach with robust variance estimation.

One of the major limitations of the present synthesis is the small number of included studies. The search yielded a few high-quality RCT in older adults that have included psychological health outcomes. Numerous studies had to be discarded due to methodological limitations or insufficient data documentation. The sample sizes of primary studies were rather small, even though this was accounted for with a bias correction factor, when pooling the data. Furthermore, we synthesized studies on older populations without a clinical diagnosis of depression or anxiety. Effects of DMI on psychological health may be more pronounced in participants with clinically relevant psychological symptoms (Alpert et al., 2009; Haboush et al., 2006; Vankova et al., 2014). Finally, due to the broad definition of DMI, the included interventions varied. Nevertheless, all types of DMI share common principles with emphasis on specific key features, which justifies synthesis of data.

### 4.8 Conclusion

This systematic evaluation of primary studies suggests that, compared to comparators, DMI improve overall psychological health in older adults without dementia. Thus, DMI may serve as a promising tool in the promotion of healthy aging and early intervention of age-related conditions. The implementation of high methodological standards and well-designed assessments of psychological health outcomes and neurophysiological mechanisms of action should be a key goal in future trials. Effective transfer of this lifestyle activity into everyday life could be expected to retain positive effects of DMI on psychological wellbeing, which needs investigation in studies of longer duration.

## Data Availability

The data that support findings of the present study are available on reasonable request from the corresponding author (OP). The R scripts necessary to reproduce the present analysis are available from the corresponding author (OP).

## 5 Acknowledgments

We would like to thank Dr. Rene Mauer (Institute for Medical Informatics and Biometry, Faculty of Medicine, Dresden University of Technology, Dresden, Germany) for the statistical consultation and support in data analysis.

## 6 Declarations

### 6.2 Authors’ contributions

OP, MW, TK: conceptualization and design of the study. OP, TW, MW, TK: methodology, statistical analysis and consultation. OP, LS: literature searches and screening, risk of bias rating, data extraction. OP, TW, SK, MW: drafting, editing and/or revision of the manuscript. OP, TW, CC, SK, MW: interpretation of results, comments on relevant literature and content. All authors contributed to the article and approved the submitted version.

### 6.3 Ethical Approval

not applicable

