## Supplement for "The impact of dance movement interventions on psychological health in older adults without dementia: A systematic review and meta-analysis"

### **Supplementary Material**

### 1 List of abbreviations and symbols

C1/C2	Outcome Cluster 1/2

DMI	Dance Movement Intervention

DMT	Dance/Movement Therapy

e.g.	for example

ENR	Enriched Environment

MesH	Medical Subject Headings

PICO	research question format: Patient – Intervention – Comparator - Outcome

PRISMA	Preferred Reporting Items for Systematic Reviews and Meta-Analyses

QoL	Quality of Life

RCT	Randomized Controlled Trial

RoB2	Risk of Bias Tool 2

RVE	Robust Variance Estimation

**Disorders**

AD	Alzheimer’s Disease

MCI	Mild Cognitive Impairment

SCD	Subjective Cognitive Decline

**Instruments and assessment tools**

36-SF	36-Short Form

BAI	Beck Anxiety Inventory

BPI	Brief Pain Inventory

GDS	Geriatric Depression Scale

GSE	General Self-Efficacy Scale

HADS-A	Hospital Anxiety and Depression Scale – Anxiety

HADS-D	Hospital Anxiety and Depression Scale – Depression

HPLP2	Health Promoting Lifestyle Profile -2

LSNS	Lubben Social Network Scale

LSS	Life Satisfaction Scale

MHC	Mental Health Continuum Short Form

MMSE	Mini-Mental State Examination

MoCA	Montreal Cognitive Assessment Test

NPI	Neuropsychiatric Inventory

PGC	Philadelphia Geriatric Center Morale Scale

PSQI	Pittsburgh Sleep Quality Index

PSS	Perceived Stress Scale

SF-12	Health related Quality of Life Short Form -12

STAI-State	State-Trait Anxiety Inventory – State

STAI-Trait	State-Trait Anxiety Inventory – Trait

SWLS	Satisfaction with Life Scale

Organizations and Institutions

ADI	Alzheimer’s Disease International

ADTA	American Dance Therapy Association

DZNE	Deutsches Zentrum für Neurodegenerative Erkrankungen e.V.

WHO	World Health Organization

### 2 PRISMA Checklist

| **Section and Topic** | **Item #** | **Checklist item** | **Location where item is reported** |
| --- | --- | --- | --- |
| **TITLE** | | |  |
| Title | 1 | Identify the report as a systematic review. | 1 |
| **ABSTRACT** | | |  |
| Abstract | 2 | See the PRISMA 2020 for Abstracts checklist. | 2 |
| **INTRODUCTION** | | |  |
| Rationale | 3 | Describe the rationale for the review in the context of existing knowledge. | 3-4 |
| Objectives | 4 | Provide an explicit statement of the objective(s) or question(s) the review addresses. | 4 |
| **METHODS** | | |  |
| Eligibility criteria | 5 | Specify the inclusion and exclusion criteria for the review and how studies were grouped for the syntheses. | 5-6 |
| Information sources | 6 | Specify all databases, registers, websites, organisations, reference lists and other sources searched or consulted to identify studies. Specify the date when each source was last searched or consulted. | 6-7; Supplement: 7 |
| Search strategy | 7 | Present the full search strategies for all databases, registers and websites, including any filters and limits used. | 7-8; Supplement: 7 |
| Selection process | 8 | Specify the methods used to decide whether a study met the inclusion criteria of the review, including how many reviewers screened each record and each report retrieved, whether they worked independently, and if applicable, details of automation tools used in the process. | 7 |
| Data collection process | 9 | Specify the methods used to collect data from reports, including how many reviewers collected data from each report, whether they worked independently, any processes for obtaining or confirming data from study investigators, and if applicable, details of automation tools used in the process. | 7 |
| Data items | 10a | List and define all outcomes for which data were sought. Specify whether all results that were compatible with each outcome domain in each study were sought (e.g. for all measures, time points, analyses), and if not, the methods used to decide which results to collect. | 8-9, Supplement: 12-13 |
|  | 10b | List and define all other variables for which data were sought (e.g. participant and intervention characteristics, funding sources). Describe any assumptions made about any missing or unclear information. | 7-8 |
| Study risk of bias assessment | 11 | Specify the methods used to assess risk of bias in the included studies, including details of the tool(s) used, how many reviewers assessed each study and whether they worked independently, and if applicable, details of automation tools used in the process. | 8 |
| Effect measures | 12 | Specify for each outcome the effect measure(s) (e.g. risk ratio, mean difference) used in the synthesis or presentation of results. | 8-9 |
| Synthesis methods | 13a | Describe the processes used to decide which studies were eligible for each synthesis (e.g. tabulating the study intervention characteristics and comparing against the planned groups for each synthesis (item #5)). | 6-7 |
|  | 13b | Describe any methods required to prepare the data for presentation or synthesis, such as handling of missing summary statistics, or data conversions. | 8-9 |
|  | 13c | Describe any methods used to tabulate or visually display results of individual studies and syntheses. | 9 |
|  | 13d | Describe any methods used to synthesize results and provide a rationale for the choice(s). If meta-analysis was performed, describe the model(s), method(s) to identify the presence and extent of statistical heterogeneity, and software package(s) used. | 8-9; Supplement: 14-15 |
|  | 13e | Describe any methods used to explore possible causes of heterogeneity among study results (e.g. subgroup analysis, meta-regression). | 9 |
|  | 13f | Describe any sensitivity analyses conducted to assess robustness of the synthesized results. | 9 |
| Reporting bias assessment | 14 | Describe any methods used to assess risk of bias due to missing results in a synthesis (arising from reporting biases). | 8 |
| Certainty assessment | 15 | Describe any methods used to assess certainty (or confidence) in the body of evidence for an outcome. | n.a. |
| **RESULTS** | | |  |
| Study selection | 16a | Describe the results of the search and selection process, from the number of records identified in the search to the number of studies included in the review, ideally using a flow diagram. | 11 |
|  | 16b | Cite studies that might appear to meet the inclusion criteria, but which were excluded, and explain why they were excluded. | n.a. |
| Study characteristics | 17 | Cite each included study and present its characteristics. | 12-15; Supplement: 9-11 |
| Risk of bias in studies | 18 | Present assessments of risk of bias for each included study. | 18-19; Supplement: 13 |
| Results of individual studies | 19 | For all outcomes, present, for each study: (a) summary statistics for each group (where appropriate) and (b) an effect estimate and its precision (e.g. confidence/credible interval), ideally using structured tables or plots. | 19-20 Supplement:  16-18 |
| Results of syntheses | 20a | For each synthesis, briefly summarise the characteristics and risk of bias among contributing studies. | 19; Supplement: 13-14 |
|  | 20b | Present results of all statistical syntheses conducted. If meta-analysis was done, present for each the summary estimate and its precision (e.g. confidence/credible interval) and measures of statistical heterogeneity. If comparing groups, describe the direction of the effect. | 19-22 |
|  | 20c | Present results of all investigations of possible causes of heterogeneity among study results. | 19-22 |
|  | 20d | Present results of all sensitivity analyses conducted to assess the robustness of the synthesized results. | 19-22 |
| Reporting biases | 21 | Present assessments of risk of bias due to missing results (arising from reporting biases) for each synthesis assessed. | 19 |
| Certainty of evidence | 22 | Present assessments of certainty (or confidence) in the body of evidence for each outcome assessed. | n.a. |
| **DISCUSSION** | | |  |
| Discussion | 23a | Provide a general interpretation of the results in the context of other evidence. | 23-25 |
|  | 23b | Discuss any limitations of the evidence included in the review. | 27 |
|  | 23c | Discuss any limitations of the review processes used. | 27 |
|  | 23d | Discuss implications of the results for practice, policy, and future research. | 26 |
| **OTHER INFORMATION** | | |  |
| Registration and protocol | 24a | Provide registration information for the review, including register name and registration number, or state that the review was not registered. | 5 |
|  | 24b | Indicate where the review protocol can be accessed, or state that a protocol was not prepared. | 5 |
|  | 24c | Describe and explain any amendments to information provided at registration or in the protocol. | n.a. |
| Support | 25 | Describe sources of financial or non-financial support for the review, and the role of the funders or sponsors in the review. | 29 |
| Competing interests | 26 | Declare any competing interests of review authors. | 29 |
| Availability of data, code and other materials | 27 | Report which of the following are publicly available and where they can be found: template data collection forms; data extracted from included studies; data used for all analyses; analytic code; any other materials used in the review. | 29 |

### 3 Search Terms

#### 3.1 Pubmed

| 1 | dance* OR dancing* OR dancing [MeSH Terms] OR dance therapy [MeSH Terms] OR “ballet” OR “jazz” OR “hiphop” OR “salsa” OR “zumba” OR “tango” OR “eurhythmics” |
| --- | --- |
| 2 | "music" AND "exercise” |
| 3 | aged [MeSH Terms] OR “aging“ OR “ageing“ OR “senior“ OR “elderly“ OR “older adults” OR “older people” OR “cognitive impairment" OR "cognitive decline" |
| 4 | psychological* OR neuropsychological* OR "mental health" OR "quality of life" OR "wellbeing" OR "life satisfaction" OR “mood” |
| 5 | random* OR control* OR “clinical trial” |
| 6 | 1 OR 2 AND 3 AND 4 AND 5 |

#### 3.2 Web of Science

| 1 | TS=(dance* OR dancing* OR dancing OR dance therapy OR “ballet” OR “jazz” OR “hiphop” OR “salsa” OR “zumba” OR “tango” OR “eurhythmics”) |
| --- | --- |
| 2 | TS=(aged OR “aging“ OR “ageing“ OR “senior“ OR “elderly“ OR “older adults” OR “older people” OR “cognitive impairment" OR "cognitive decline") |
| 3 | TS=(psychological* OR neuropsychological* OR "mental health" OR "quality of life" OR "wellbeing" OR "life satisfaction" OR “mood”) |
| 4 | TS=(random* OR control* OR “clinical trial”) |
| 5 | 1 AND 2 AND 3 AND 5 |

#### 3.3 PsycInfo (via EBSCO)

| 1 | dance* OR dancing* OR dancing OR dance therapy [MeSH Terms] OR “ballet” OR “jazz” OR “hiphop” OR “salsa” OR “zumba” OR “tango” OR “eurhythmics” |
| --- | --- |
| 2 | "music" AND "exercise” |
| 3 | aged [MeSH Terms] OR “aging“ OR “ageing“ OR “senior“ OR “elderly“ OR “older adults” OR “older people” OR “cognitive impairment" OR "cognitive decline" |
| 4 | psychological* OR neuropsychological* OR "mental health" OR "quality of life" OR "wellbeing" OR "life satisfaction" OR “mood” |
| 5 | random* OR control* OR “clinical trial” |
| 6 | 1 OR 2 AND 3 AND 4 AND 5 |

### 4 Characteristics of interventions

**Table S1.** Characteristics of Dance Movement Interventions (DMI) and Control Conditions.

| **Study** |  | **DMI** | | | | | | | |  | **Control Intervention** | |
| --- | --- | --- | --- | --- | --- | --- | --- | --- | --- | --- | --- | --- |
|  | **Type of DMI** | | **Description** | **Period and Frequencies** | **Qualification of instructor** | **Manual** | **Setting** | **Type of Music/ Rhythm** | **Adherence** |  | **Control Type(s)** | **Description** |
| Alves, 2013 | Ballroom Dance | | Groups learned dance sequences of different rhythms led by certified instructor; warm-up and cool-down | 16 weeks, 2/week, 120 min | Certified dance instructor | NR | group setting | e.g. Samba, Bolero | 90% |  | AC: Walking  PC: no contact | Walking in a group matched to activity level of DMI group, passive control |
| Bisbe et al., 2020 | Choreo-graphy | | Performing aerobic dances in groups of 8 in light to moderate intensity led by physical therapist and videos for visual support | 12 weeks, 2/week, 60 min | Physical Therapist specialzed in geriatrics | yes | Day care hospital, group setting | Salsa, Rock, Jive, Pop | 95% |  | AC: Physical Therapy | Multimodal physical therapy program (strength, gait, flexibility, balance training) |
| Chang et al., 2021 | Square Dance | | Chinese square dance performed outdoors in groups with practice videos before intervention for familiarization | 18 weeks, 3/week, 30 min | National social sports instructors | NR | Group setting, outdoor | Dance music with simple melodies | 88% |  | PC: Usual Care | Liberal daily lifestyle |
| Cruz-Ferreira et al., 2015 | Creative Dance | | Five elements of movement: body, space, time, dynamic and relationship, performed in groups | 24 weeks, 2/week, 50 min | Nurse, dance teacher | NR | Local health center, group setting | differed to theme (e.g. classic, jazz, pop, ethnics) | 85% |  | PC: Waitlist | NA |
| Esmail et al., 2020 | Dance Movement | | ADTA-based program adapted to older adults | 12 weeks, 3/week, 60 min | Registered ADTA therapist | Yes | Gym facility, group setting | different styles | 91% |  | AC: Aerobic Exercise Training  PC: Waitlist | Cardio-vascular training on bicycle |
| Eyigor et al., 2009 | Folkloric Dance | | Dancing folkloric routines in groups led by dance senior, additional instruction to walk for 30 minutes twice a week | 8 weeks, 3/week, 60 min | folklore dance expert | NR | Reha-bilitation unit, group setting | Turkish folk music | NR |  | PC: no intervention | NA |
| Hars et al., 2014 | Eurythmy | | Multitask exercises of progressive difficulty with different objectives in a group | 25 weeks, 1/week, 60 min | Experienced instructor | Yes | group setting | Piano music | 79% |  | PC: Waitlist | NA |
| Hui et al., 2009 | Aerobic Dance | | Low impact dancing choreography consisting of cross steps and cha-cha steps in a group created by professional dance instructor and physical therapist | 12 weeks, 2/week, 50 min | Professional dance instructor, physical therapist | NR | group setting | Canto-Pop Song | 92% |  | PC: no intervention | NA |
| Kosmat & Vranic, 2017 | Standard Dance | | dancing/ learning slow waltz under supervision of trained dance pedagogist | 10 weeks, 1/week, 45 min | Trained dance teacher | NR | Care center, group setting | Waltz | NR |  | AC: Social Discussion | Various topics (needs and interests of participants) discussed in groups |
| Lazarou et al., 2017 | Ballroom Dance | | Learning several ballroom dances with routines and music led by experienced dance instructor | 40 weeks, 2/week, 60 min | Dance instructor | NR | group setting | Tango, Waltz, Rumba, Chachacha, Swing Salsa, etc. | NR |  | PC: no intervention | NA |
| Liao et al., 2018 | Music and Tai-Chi (Combined) | | Combination of 24 Tai Chi movements and music in groups | 12 weeks, 3/week, 50 min | NR | NR | group setting | Soft Chinese folk music | NR |  | AC: Routine health education | NR |
| Serrano-Guzmán et al., 2016 | Dance Therapy (Flamenco) | | Low-impact aerobics and stretching mixed with dance movements based on flamenco performed in groups | 8 weeks, 3/week, 50 min | NR | Yes | group setting | Flamenco | 100% |  | AC: Self-care treatment advice | Follow physical activity recom-mendations |
| Zhu et al., 2018 | Aerobic Dance | | learning dance routine in groups together with dance instructor | 12 weeks, 3/week, 35 min | Dance instructor, physical therapist | NR | group setting | Musical phrases | median attended sessions: 36 |  | PC: no intervention | NA |

*Note.* DMI, Dance Movement Intervention; AC, Active Control; PC, Passive Control; NR, Not reported; NA, Not applicable, ADTA, American Dance Therapy Association.

### 5 Incorporated Outcome Measures

#### 5.1 Positive psychological outcomes

**Table S2.** Assessment tools for psychological health in the positive domain.

| **Name of Tool** | **Subscale (if applicable)** | **Psychological Construct** | **k** |
| --- | --- | --- | --- |
| General Self Efficacy Scale (GSE) | - | Quality of Life | 1 |
| Health Promoting Lifestyle Profile II (HPLP-II) | - | Quality of Life | 1 |
| Life Satisfaction Scale | - | Quality of Life | 1 |
| Lubben Social Network Scale (LSNS) | - | Social Integration | 1 |
| Mental Health Continuum Short Form (MHC) | - | Well-Being | 1 |
| Pittsburgh Sleep Quality Index (PSQI) | - | Quality of Life | 1 |
| Ryff’s Psychological Well-Being Scales | - Autonomy - Environmental Mastery - Personal Growth - Positive Relations with others - Purpose in Life - Self-Acceptance | (subjective) Well-Being | 1 |
| Satisfaction with Life Scale (SWLS) | - | Quality of Life | 1 |
| SF-12 Health related quality of life | - Mental Component | Quality of Life | 3 |
| SF-36 | - General Health - Physical Functioning - Role – Physical - Pain - Vitality - Mental Health - Role – Emotional - Social Functioning | Quality of Life | 2 |

*Note.* k, number of studies using the tool; SF-12, Short-Form-12; SF-36, Short-Form-36.

#### 5.2 Negative psychological outcomes

**Table S3**. *Assessment tools for psychological health in the negative domain.*

| **Name of Tool** | **Subscale (if applicable)** | **Psychological Construct** | **k** |
| --- | --- | --- | --- |
| Beck Anxiety Inventory (BAI) | - | Anxiety | 1 |
| Brief Pain Inventory (BPI) | - | Pain | 1 |
| Geriatric Depression Scale (GDS and GDS-15) | - | Depression | 4* |
| Hospital Anxiety and Depression Scale (HADS) | - HADS-A - HADS-D | Anxiety  Depression | 2 |
| Neuropsychiatric Inventory (NPI) | - | Neuropsychiatric Burden | 1 |
| Perceived Stress Scale (PSS) | - | Stress | 1 |
| State-Trait Anxiety Inventory (STAI) | - STAI-State - STAI-Trait | Anxiety | 1 |

*Note.* k, number of studies using the tool. *Includes the study by Eyigor et al., 2009, where GDS was assessed but no data could be extracted as the outcome was reported as percentage and was therefore not included in quantitative synthesis.

#### 5.3 Additional outcomes

**Table S4**. Assessment tools for secondary outcome cognitive function.

| **Name of Tool** | **Psychological Construct** | **k** |
| --- | --- | --- |
| Mini Mental State Examination (MMSE) | General Cognitive Abilities | 3 |
| Montréal Cognitive Assessment (MoCA) | General Cognitive Abilities | 4 |
| Raven’s Advanced Matrices | General Cognitive Abilities | 1 |

*Note.* k, number of studies using the tool.

### 6 Risk of Bias Rating

Methodological quality of each study was assessed with the Risk of Bias Tool 2 (1). Resulting ratings for individual studies according to domains assessed can be seen in figure S1. Each study was rated regarding an intention-to-treat approach. Detailed information on each domain is given below.

**Figure S1.** Risk of Bias rating for individual studies.

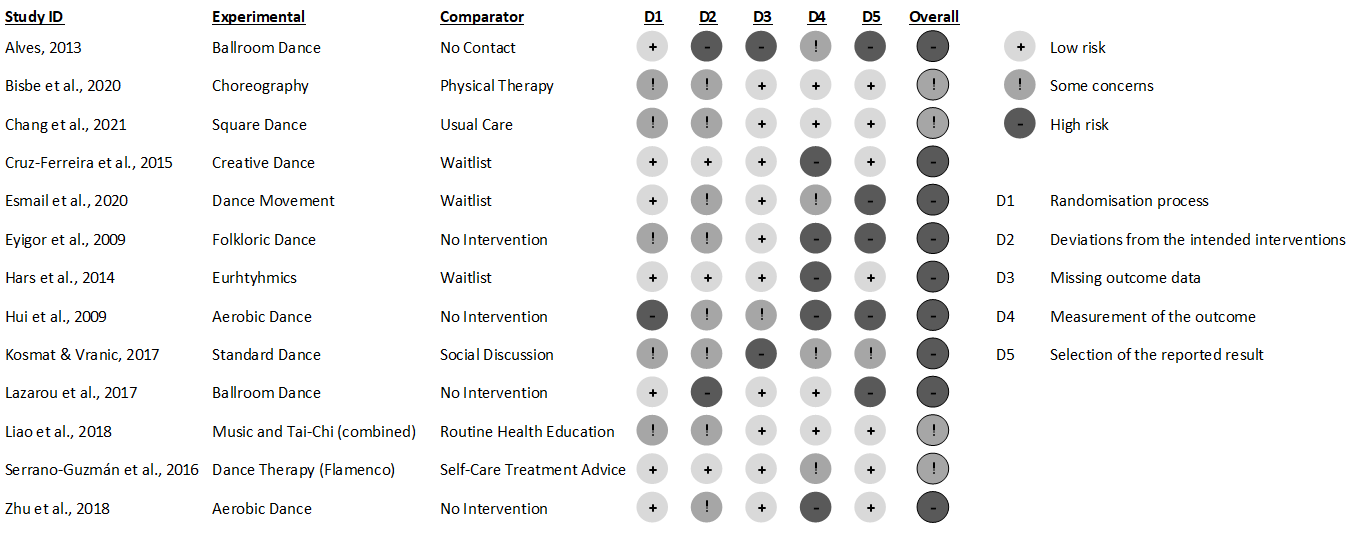

*Note.* Risk of bias was assessed with the RoB tool 2 regarding an intention-to-treat methodology following the five domains (D1-D5).

The rating procedures yielded following results:

- **“Randomization process” (D1) bias rating:** A large proportion of studies was rated as being at “low risk” (*k* = 7, 54%) or “some concerns” (*k* = 5, 38%). Individual studies confirmed a successful randomization process by showing no significant differences in baseline measures between groups.
- “**Deviations from the intended interventions” (D2) bias rating:** Eight studies (62%) were rated with “some concerns” and two studies (15%) were rated at “high risk”. “High” bias ratings reflected the use of per-protocol analysis (*k* = 8; 62%) and bias for blinding (*k* = 4, 31%) as inherent to non-pharmacological interventions. Blinding of participants is barely possible due to the nature of DMI. Blinding of study assessors was reported in four studies (31%). Psychological outcomes were assessed via patient/participant-reported outcomes, which could be biased. Three studies (23%) applied an intention-to-treat analysis and were rated at “low risk”.
- **“Missing outcome data” (D3) bias rating:** A large proportion of studies (*k* = 10, 77%) was assigned to “low risk” of bias. One study reported a high drop-out rate for only one group which resulted in an unbalanced group size included in the statistical analyses (2) and was therefore rated as being at high risk. The remaining two studies did not provide sufficient information on how missing outcome data occurred and was handled.
- **“Measurement of the outcome” (D4) bias rating:** Four studies (31%) were rated with “some concerns” and five studies (38%) were rated at “high risk”. Risk of bias was seen in the self-reported assessment of psychological health and participants’ awareness of group allocation especially with passive controls.
- **“Selection of reported outcomes” (D5) bias rating:** Ratings were split between studies rated at “low risk” (*k* =7, 54%) or studies rated as “high risk” (*k* =5, 38%). Risk of bias was seen for studies that reported selected psychological health outcomes in the result sections.

### 7 Statistical analyses and formulas

The calculations described below were applied and implemented using the free statistical computation software R (version 4.0.3) for each outcome score to determine effect sizes. Firstly, the standardized mean change within each study arm (DMI and control condition) between pre- to post-intervention scores was calculated with the “metafor” package (version 3.0-2) in R (3). Secondly, the difference between these change scores and sampling variance was determined. Following Morris (4) this score was then divided by the pooled pre-test standard deviations and a bias correction factor was applied resulting in the effect size g (formula given below). The formula of the present effect size calculation involves knowledge on pre-test post-test correlations, which are rarely reported. A value of r = 0.50 was employed as a common value for substitution of unknown correlations (5, 6).

Effect size formula (see equation 8 in Morris (4)):

$$d_{ppc2}=c_{P} \left[ \frac{\left( M_{post,T}-M_{pre,T} \right)-(M_{post,C}-M_{pre,C})}{{SD}_{pre}} \right]$$

with the pooled standard deviation (see equation 9):

$${SD}_{pre}= \sqrt{\frac{\left( n_{T}-1 \right){SD}_{pre,T}^{2}+(n_{C}-1){SD}_{pre,C}^{2}}{n_{T}-n_{C}-2}}$$

and the bias correction factor was defined according to equation 6 e in Hedges (7)

$$c\left( m \right)= \frac{\Gamma\left( \frac{m}{2} \right)}{\sqrt{\frac{m}{2}} \Gamma\left( \frac{m-1}{2} \right)}$$

with:	$m=n_{T}- n_{C}-2$

the variance of effect size was calculated according to equation 13 in Becker (8)

$$var=2\left( 1- r \right)\left( \frac{1}{n_{T}}+\frac{1}{n_{C}} \right)+\frac{\Delta^{2}}{2\left( n_{T}+ n_{C} \right)}$$

| **Operator** | **Definition** |
| --- | --- |
| *M* | Mean score of a test outcome |
| *T* | Treatment group |
| *C* | Control group |
| *Post* | Immediately post-intervention |
| *Pre* | prior to intervention (baseline assessment) |
| *n* | Sample size of group |
| *r* | Correlation between M_pre_ and M_post_ |
| *C_p_* | Bias correction factor |
| *SD* | Standard Deviation |
| △ | Effect Size |

### 8 Publication Bias

**Figure S2.** Funnel Plot.

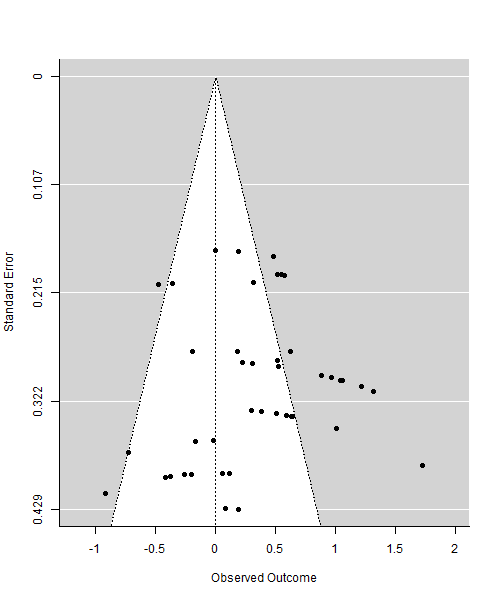

*Note.* Observed effect sizes per individual outcome measure included in data synthesis were plotted against standard error. Assessment of asymmetry was confirmed with an Egger’s regression test indicating no evidence for publication bias.

### 9 Forest plots

#### 9.1 Positive psychological domain

**Figure S3.** Forest plot of effect sizes of DMI compared to passive control type (where applicable otherwise active control groups were used as comparator) on the positive outcome domain for individual studies and outcome measures.

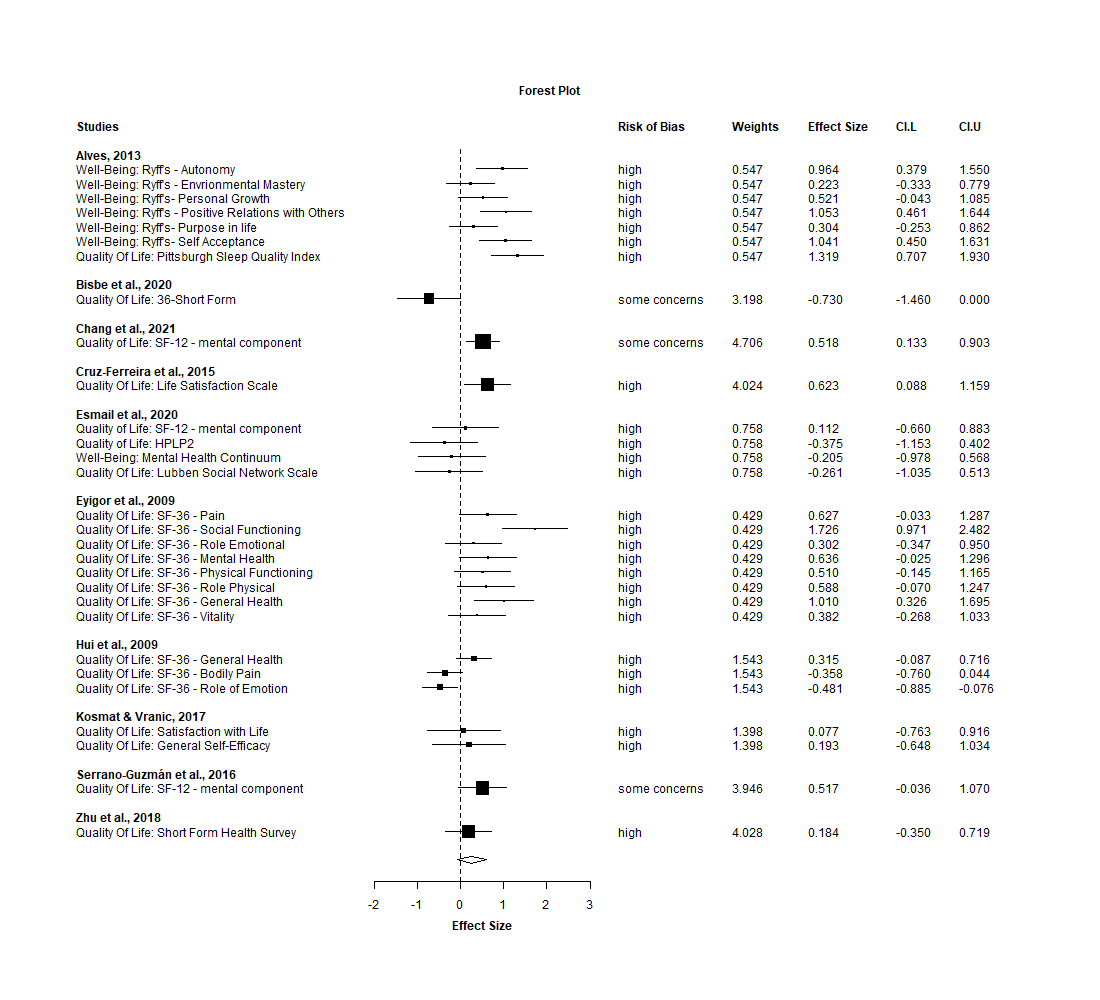
*Note.* Effect sizes are presented for each study according to constructs in the positive outcome domain (psychological health) and assessment tools used, respectively. Values on the left side of the null line favor control condition and values on the right side favor DMI. **Key**: CI.L, Confidence interval lower bound; CI.U, Confidence interval upper bound.

#### 9.2 Negative psychological domain

**Figure S4.** Forest plot of effect sizes of DMI compared to passive control type (where applicable otherwise active control groups were used as comparator) on the negative outcome domain for individual studies and outcome measures.

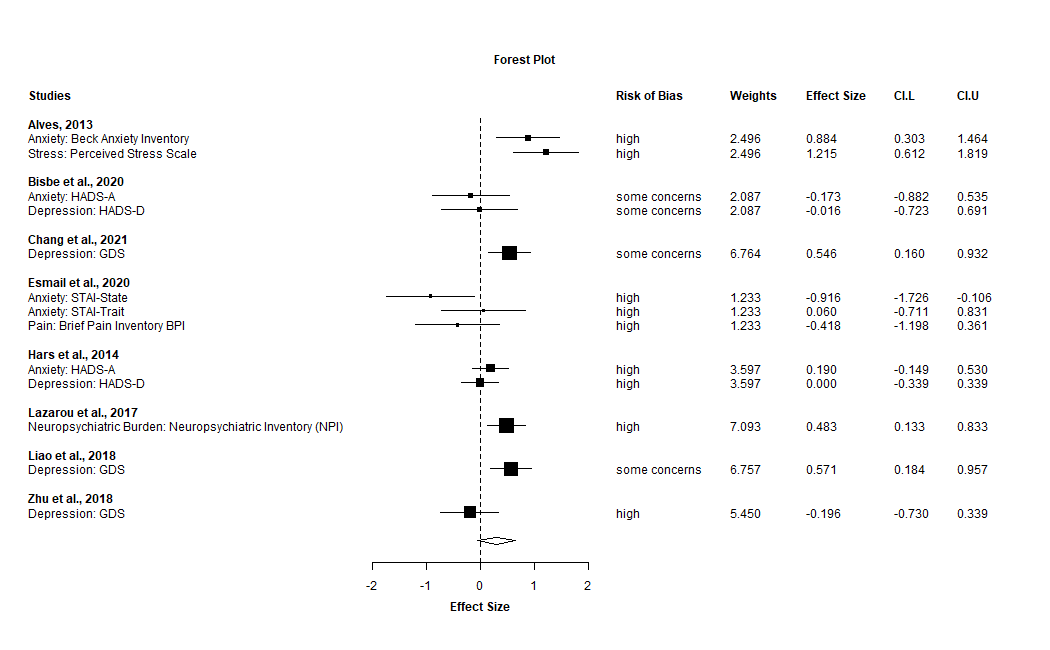

*Note.* Effect sizes are presented for each study according to constructs in the negative outcome domain (psychological health) and assessment tools used, respectively. Values on the left side of the null line favor control condition and values on the right side favor DMI. ***Key*:** CI.L, Confidence interval lower bound; CI.U, Confidence interval upper bound; HADS-A/D, Hospital Anxiety and Depression Inventory; GDS, Geriatric Depression Scale.

#### 9.3 General cognitive function

**Figure S5.** Forest Plot of effect sizes of DMI compared to passive control type (where applicable otherwise active control groups were used as comparator) on general cognitive abilities for individual studies and outcome measures.

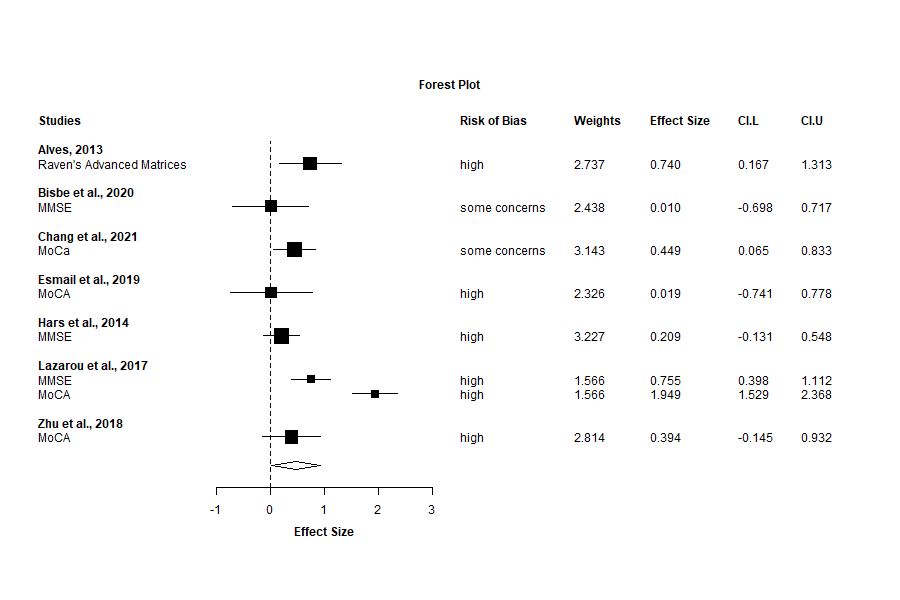
*Note.* Effect sizes are presented for each study according to measurement tools of general cognitive abilities, respectively. Values on the left side of the null line favor control condition and values on the right side favor DMI. ***Key*:** CI.L, Confidence interval lower bound; CI.U, Confidence interval upper bound; MMSE, Mini-Mental State Examination; MOCA, Montreal Cognitive Assessment.

### 10 Outlier Analysis

Post-hoc inspection of studies with regard to possible outliers resulted in the identification of six individual scales. Following scales were excluded as their particular CIs did not overlap with the overall effect CI:

- Pittsburgh Sleep Quality Index; Perceived Stress Scale (2)
- 36-Short Form (9)
- STAI-State (10)
- SF-36 social functioning (11)
- SF-36 Bodily Pain; SF-36 Role of Emotion (12)

Analyses were repeated as described in the method section with results provided below (see Table S5). Lastly, exclusion of the study by Hui and colleagues (12), due to methodological considerations concerning the randomization process, substantiated our findings for overall psychological health (g = 0.37; CI 95% [0.15, 0.58]; p = 0.003; I² = 51.66) and the positive domain (g = 0.33; CI 95% [-0.01, 0.67]; p = 0.06; I² = 58.88).

**Table S5.** Meta-analyses comparing DMI to passive control type (where applicable) with outliers removed for psychological health outcomes.

|  | **Construct** | **k *(N* ES)** | **ES** | **95% CI** | **SE** | **df** | ***p-*value** | **I² %** |
| --- | --- | --- | --- | --- | --- | --- | --- | --- |
| Overall  psychological health | all combined | 13(35) | 0.37 | [0.21, 0.53] | 0.07 | 10 | .0005 | 25.39 |
| Positive domain | all combined | 9(24) | 0.42 | [0.25, 0.58] | 0.07 | 6 | .0007 | 0 |
|  | Quality of life | 8(17) | 0.39 | [0.21, 0.57] | 0.07 | 5 | .002 | 0 |
| Negative domain | all combined | 8(11) | 0.31 | [0.01, 0.62] | 0.13 | 6 | .05 | 56.64 |
|  | Anxiety | 4(4) | 0.27 | [-0.44, 0.97] | 0.21 | 3* | * | 52.28 |
|  | Depression | 5(5) | 0.22 | [-0.25, 0.68] | 0.16 | 4 | .26 | 60.81 |
| ***Key.*** DMI, Dance Movement Intervention; k, number of studies; *N* ES, Number of Effect Sizes; ES, Effect size; CI, Confidence interval; SE, Standard Error; df, Degrees of freedom.  *Where df <4, p-values are unreliable and are thus not reported here. | | | | | | | | |
